# A Machine Learning Approach to Prediction and Multimorbidity Risk Factor Identification in a low- and middle-income country

**DOI:** 10.1101/2025.10.13.25337900

**Authors:** Olalekan A. Uthman, Matthew Hazell, Muhammed Mubashir Babatunde Uthman, Kolawole W Wahab, Ponnusamy Saravanan, Paramjit Gill, Andre Pascal Kengne

**Author notes:** **Corresponding author:** Olalekan A. Uthman. Warwick Applied Health, Warwick Medical School, University of Warwick, Coventry, CV4 7AL, United Kingdom.

## Abstract

**Importance:** Multimorbidity, the coexistence of multiple chronic conditions, is a growing public health challenge, particularly in low- and middle-income countries like South Africa. Identifying individuals at high risk of multimorbidity is crucial for developing targeted interventions and allocating healthcare resources effectively.

**Objective:** To investigate the predictive performance of various machine learning models in identifying individuals at risk of multimorbidity in South Africa and to identify the most influential predictors of multimorbidity, considering both individual-level and contextual factors.

**Design, Setting, and Participants:** This cross-sectional study utilized data from the South Africa Demographic and Health Survey (SADHS) 2016, a nationally representative household survey. The study included 5,342 participants aged 18 years and older, of which 2,107 (33.9%) had multimorbidity, defined as the presence of two or more chronic conditions.

**Main Outcomes and Measures:** The primary outcome was the presence of multimorbidity. Machine learning models, including gradient boosting classifier, linear discriminant analysis, ada boost classifier, logistic regression, ridge classifier, catboost classifier, random forest classifier, light gradient boosting machine, extra trees classifier, naive bayes, quadratic discriminant analysis, extreme gradient boosting, k neighbors classifier, dummy classifier, decision tree classifier, svm - linear kernel, were developed and evaluated using a repeated train-test split approach. Model performance was assessed using accuracy, area under the receiver operating characteristic curve (AUC), recall, precision, F1 score, Cohen’s Kappa, and Matthews Correlation Coefficient (MCC). Shapley Additive Explanations (SHAP) were used to identify the most influential predictors of multimorbidity.

**Results:** The Gradient Boosting Classifier achieved the highest predictive performance, with an AUC of 0.7809, accuracy of 0.7478, and F1 score of 0.5798. Age, no medication use, sex, poor health perception, and community illiteracy rate were identified as the most influential predictors of multimorbidity. Individual-level factors had a more substantial impact on the likelihood of multimorbidity compared to community-level factors. However, higher community illiteracy rates and regional unemployment rates were associated with an increased likelihood of multimorbidity, highlighting the importance of contextual factors. The fairness and demographic bias assessment revealed that the Gradient Boosting Classifier maintained a high level of fairness across different regions, wealth index categories, age groups, and genders.

**Conclusion and Relevance:** Machine learning algorithms, particularly the Gradient Boosting Classifier, can accurately predict multimorbidity in the South African context. The findings emphasize the importance of considering both individual-level and contextual factors in understanding the determinants of multimorbidity.

## Background

Multimorbidity, defined as the co-occurrence of multiple long-term conditions within an individual, has emerged as a significant global health challenge, disproportionately affecting low- and middle-income countries (LMICs) (Afshar et al., 2015). In South Africa, a country grappling with a double burden of communicable and non-communicable diseases, the prevalence of multimorbidity is on the rise (Weimann et al., 2016). The complex interplay of socioeconomic determinants, including neighbourhood-level factors, has been recognized as a crucial driver of this growing burden (Hurst et al., 2015).

The impact of multimorbidity on individuals, healthcare systems, and societies in LMICs is profound. Patients with multiple chronic conditions often experience decreased quality of life, functional limitations, and increased healthcare utilization (Arokiasamy et al., 2015). Healthcare systems in LMICs, already strained by resource constraints, face the challenge of providing comprehensive and coordinated care for individuals with complex health needs (Oni et al., 2015). Moreover, the economic burden of multimorbidity, both in terms of direct healthcare costs and indirect costs such as lost productivity, poses a significant threat to the socioeconomic development of these nations (Wang et al., 2014).

Recognising the urgent need to address the growing burden of multimorbidity in LMICs, researchers have sought to identify the underlying determinants and predictors of this phenomenon. While individual-level factors such as age, gender, and lifestyle behaviours have been extensively studied, the role of neighbourhood-level socioeconomic factors in shaping the risk of multimorbidity has garnered increasing attention (Alaba & Chola, 2013). Neighbourhood socioeconomic status (SES), encompassing factors such as income levels, educational attainment, and employment rates, has been linked to the development and progression of chronic diseases (Weimann et al., 2016).

Recent advancements in machine learning have opened up new avenues for exploring the complex relationships between neighbourhood SES and multimorbidity in LMICs (Panch et al., 2018). By leveraging large datasets and advanced analytical techniques, machine learning models can uncover intricate patterns and predictors that traditional statistical methods may overlook (Prosperi et al., 2020). This approach holds immense potential for identifying neighbourhood-level socioeconomic determinants of multimorbidity, enabling the development of targeted interventions and policies to mitigate the burden of this condition in resource-limited settings.

Although the influence of neighbourhood socioeconomic status (SES) on health outcomes is increasingly acknowledged, to the best of our knowledge, machine learning approaches have not been employed to examine its predictive utility for multimorbidity within low- and middle-income country (LMIC) settings such as South Africa. This study aims to address this gap by harnessing the power of machine learning to elucidate the relationship between neighbourhood socioeconomic indicators and multimorbidity.

## Methods

### Study Design and Data Sources

This study employed a cross-sectional design using data from the South Africa Demographic and Health Survey (SADHS) 2016. The SADHS 2016 is a nationally representative household survey that provides comprehensive information on various demographic and health indicators (NDoH et al., 2019). The DHS programme has assisted in conducting over 350 nationally representative household surveys across 90 countries since 1984, making it an essential data source for policy-making, monitoring, and evaluation in many LMICs (Corsi et al., 2012; USAID, 2023).

The SADHS 2016 utilized a stratified two-stage sample design (NDoH et al., 2019). In the first stage, 750 primary sampling units (PSUs) were selected from 26 sampling strata based on urban, traditional, and rural areas within each of the nine South African provinces. In the second stage, a fixed number of 20 dwelling units (DUs) were randomly selected from each PSU. This design allows for the estimation of key variables at the national, provincial, and area (urban, rural, traditional) levels. Data collection took place from 27 June 2016 to 4 November 2016.

All DUs were eligible for the primary modules on women, fertility, and children, while half were subsampled for modules on men and adult health. The adult health module included self-reported chronic conditions and biomarker collection for anthropometry, anaemia, hypertension, HbA1c levels for diabetes, and HIV for participants aged over 15.

### Study Population

Men and women aged 18 and over were included in this study if they contributed data to the SADHS 2016 adult health modules. Individuals under 18 were excluded to ensure comparability with the literature. Participants were also excluded if they were missing information on the multimorbidity outcome.

### Variables

#### Outcome Variable

Multimorbidity was defined as the presence of two or more co-existing chronic conditions (Johnston et al., 2019). Twelve current chronic diseases were considered: tuberculosis, hypertension, stroke, high blood cholesterol, anaemia, chronic bronchitis, diabetes, asthma, cancer, heart disease, human immunodeficiency virus (HIV), and chronic pain. These conditions were recorded through self-reports and biomarker measurements. Participants with missing information on a disease (self-report or biomarker) were coded as missing rather than not having the disease to reduce misclassification bias.

#### Explanatory Variables

Explanatory variables were selected based on literature review and a-priori reasoning. Socioeconomic variables included household wealth, education level, occupational status, health insurance, and marital status. Individual-level health variables included BMI, dietary health, sugary drink intake, smoking status, alcohol consumption, and occupational smoke exposure. Access to old and new media were included as proxies for health information access. Age, sex, and ethnicity were also included. Further details on variable derivation and coding are provided in Table S1 (Supplementary file).

Neighbourhoods were defined as clusters of households serving as PSUs within the DHS. Neighbourhood and province-level explanatory variables included poverty, rurality, unemployment level, and illiteracy (community-level only). These were categorized as low, medium, or high based on the proportion of individuals in the most deprived group for each neighbourhood and province, calculated using the larger adult-health sample (N=10,336) to include more contextual information.

#### Data Preprocessing and Analysis

Data preprocessing involved cleaning, recoding variables, and handling missing data as described in the Variables section. Participants with missing information on the multimorbidity outcome were excluded from the analysis. Descriptive statistics, including means, standard deviations, and proportions, were calculated for all variables, stratified by multimorbidity status. These summary statistics provided an overview of the characteristics of the study population and potential differences between those with and without multimorbidity.

#### Machine Learning Model Development

A repeated train-test split approach was employed for model development and evaluation. The dataset was randomly divided into training (80%) and test (20%) sets, ensuring that data points from the same patient were exclusively included in either the training or test set to prevent data leakage. Various machine learning algorithms, including Gradient Boosting Classifier, Linear Discriminant Analysis, AdaBoost Classifier, Logistic Regression, Ridge Classifier, CatBoost Classifier, Random Forest Classifier, Light Gradient Boosting Machine, Extra Trees Classifier, Naive Bayes, Quadratic Discriminant Analysis, Extreme Gradient Boosting, K-Nearest Neighbors Classifier, Dummy Classifier, Decision Tree Classifier, and Support Vector Machine with Linear Kernel, were applied.

Hyperparameters for each model were optimized using random search with 10-fold cross-validation on the training set. The process of splitting, hyperparameter optimization, model training, and evaluation was repeated 1,000 times to ensure the robustness of the results and to account for potential variations due to random splitting. The performance metrics, including accuracy, AUC, recall, precision, F1 score, Cohen’s Kappa, and Matthews Correlation Coefficient (MCC), were calculated and averaged across all iterations.

#### Model Interpretation

To gain insights into the factors contributing to the prediction of multimorbidity, Shapley Additive Explanations (SHAP) were employed. SHAP values were calculated for the best-performing model, which was the Gradient Boosting Classifier with the highest AUC on the test set across all iterations. The SHAP values provided a unified measure of feature importance by quantifying the contribution of each predictor variable to the model output. The SHAP summary plot displayed the average impact of each feature on the model output, with positive values indicating an increase in the likelihood of multimorbidity and negative values indicating a decrease.

Additionally, SHAP dependence plots were generated to visualize the impact of individual-level and community-level features on the model output. These plots showed the relationship between the feature values and the SHAP values, providing a more detailed understanding of how each feature influenced the prediction of multimorbidity. The interpretation of these plots offered insights into the complex interplay between individual and contextual factors in shaping the risk of multiple chronic conditions. The SHAP analysis was conducted using Python (version 3.10.4) and the SHAP library.

#### Evaluation

The performance of the machine learning models was evaluated using various metrics, including accuracy, AUC, recall, precision, F1 score, Cohen’s Kappa, and MCC. The primary evaluation metric was the AUC, which measures the model’s ability to discriminate between individuals with and without multimorbidity. The AUC ranges from 0 to 1, with a value of 0.5 indicating a random classifier and a value of 1 indicating a perfect classifier. The other metrics provided additional insights into the model’s performance, such as the balance between precision and recall (F1 score), the agreement between predicted and actual labels (Cohen’s Kappa), and the correlation between predictions and observations (MCC).

#### Fairness and Demographic Bias Assessment

To ensure the machine learning model’s effectiveness and equitable application across diverse populations, a fairness and demographic bias assessment was integrated into the study. During the model development phase, key demographic factors, such as age, gender, region, and wealth index, were considered to capture the wide-ranging realities and experiences of the patient population. The model’s performance was evaluated across these demographic groups using various metrics, including accuracy, precision, recall, F1 score, Selection Rate, Cohen’s Kappa, and Matthews Correlation Coefficient (MCC).

## Results

### Descriptive Statistics

As presented in Table 1, the study included a total of 5,342 participants, of which 2382 (44.6%) were classified as having multimorbidity, defined as the presence of two or more chronic conditions. The mean age of participants with multimorbidity was nearly 14 years higher than those without multimorbidity. Access to old and new media, health insurance coverage, and poverty status were found to be similar between the two groups.

**Table 1:** Summary characteristics of included participants. Characteristics of the study population by multimorbidity status.

|  |  | Overall | People without multimorbidity | People with multimorbidity |
| --- | --- | --- | --- | --- |
| Total, N (% row) |  | 5,342 (100.0%) | 2,960 (55.4%) | 2,382 (44.6%) |
| *Individual socio-economic variables* |  |  |  |  |
| Wealth index, N (% col) | Poorest | 1,128 (21.1%) | 628 (21.2%) | 500 (21.0%) |
|  | Poorer | 1,063 (19.9%) | 588 (19.9%) | 475 (19.9%) |
|  | Middle | 1,150 (21.5%) | 628 (21.2%) | 521 (21.9%) |
|  | Richer | 1,039 (19.5%) | 569 (19.2%) | 470 (19.7%) |
|  | Richest | 962 (18.0%) | 546 (18.4%) | 416 (17.5%) |
| Education, N (% col) | No education | 448 (8.4%) | 154 (5.2%) | 294 (12.3%) |
|  | Primary | 1,021 (19.1%) | 437 (14.8%) | 584 (24.5%) |
|  | Secondary | 3,342 (62.6%) | 2,018 (68.2%) | 1,324 (55.6%) |
|  | Higher | 531 (9.9%) | 351 (11.8%) | 180 (7.6%) |
| Occupation, N (% col) | Unemployed | 3,464 (64.9%) | 1,856 (62.7%) | 1,609 (67.5%) |
|  | Professional/technical | 304 (5.7%) | 195 (6.6%) | 108 (4.6%) |
|  | Clerical | 166 (3.1%) | 102 (3.4%) | 65 (2.7%) |
|  | Agricultural | 69 (1.3%) | 40 (1.3%) | 29 (1.2%) |
|  | Domestic service | 100 (1.9%) | 46 (1.6%) | 54 (2.3%) |
|  | Sales and services | 274 (5.1%) | 161 (5.4%) | 113 (4.7%) |
|  | Skilled manual | 360 (6.7%) | 246 (8.3%) | 114 (4.8%) |
|  | Unskilled manual | 399 (7.5%) | 177 (6.0%) | 222 (9.3%) |
|  | Missing | 205 (3.8%) | 137 (4.6%) | 68 (2.8%) |
| Have health insurance, N (% col) | Yes | 486 (9.1%) | 321 (10.8%) | 165 (6.9%) |
| *Individual socio-demographic variables* |  |  |  |  |
| Mean age, years (SD) |  | 41.6 (17.6) | 35.5 (15.7) | 49.1 (17.0) |
| Sex, N (% col) | Men | 1,973 (36.9%) | 1,321 (44.6%) | 652 (27.4%) |
| Ethnicity, N (% col) | Black | 4,613 (86.4%) | 2,543 (85.9%) | 2,069 (86.9%) |
|  | White | 278 (5.2%) | 172 (5.8%) | 106 (4.5%) |
|  | Mixed ancestry | 378 (7.1%) | 210 (7.1%) | 169 (7.1%) |
|  | Other | 72 (1.4%) | 34 (1.2%) | 38 (1.6%) |
| Marital status, N (% col) | Never married | 2,503 (46.9%) | 1,658 (56.0%) | 845 (35.5%) |
|  | Formerly married | 711 (13.3%) | 217 (7.3%) | 494 (20.7%) |
|  | Currently married | 2,128 (39.8%) | 1,085 (36.7%) | 1,043 (43.8%) |
| Old media access, N (% col) | Low | 2,091 (39.1%) | 1,094 (37.0%) | 997 (41.8%) |
|  | Medium | 1,931 (36.1%) | 1,090 (36.8%) | 841 (35.3%) |
|  | High | 1,320 (24.7%) | 775 (26.2%) | 545 (22.9%) |
| New media access, N (% col) | Low | 545 (10.2%) | 263 (8.9%) | 282 (11.8%) |
|  | Medium | 3,001 (56.2%) | 1,420 (48.0%) | 1,581 (66.4%) |
|  | High | 1,796 (33.6%) | 1,277 (43.1%) | 519 (21.8%) |
| *Individual health variables* |  |  |  |  |
| Body mass index, N (% col) | Underweight | 222 (4.2%) | 151 (5.1%) | 72 (3.0%) |
|  | Healthy weight | 1,969 (36.9%) | 1,304 (44.0%) | 665 (27.9%) |
|  | Overweight | 1,364 (25.5%) | 743 (25.1%) | 620 (26.0%) |
|  | Obese | 1,642 (30.7%) | 668 (22.6%) | 974 (40.9%) |
|  | Missing | 144 (2.7%) | 94 (3.2%) | 50 (2.1%) |
| Smoking status, N (% col) | Never smoker | 4,051 (75.8%) | 2,165 (73.1%) | 1,887 (79.2%) |
|  | Current smoker | 1,043 (19.5%) | 659 (22.3%) | 384 (16.1%) |
|  | Former smoker | 248 (4.6%) | 136 (4.6%) | 111 (4.7%) |
| Drink alcohol, N (% col) | Yes | 1,779 (33.3%) | 1,144 (38.6%) | 635 (26.7%) |
| Dietary health, N (% col) | High | 2,533 (47.4%) | 1,287 (43.5%) | 1,246 (52.3%) |
|  | Medium | 1,648 (30.8%) | 924 (31.2%) | 724 (30.4%) |
|  | Poor | 1,161 (21.7%) | 749 (25.3%) | 412 (17.3%) |
| Sugary drink intake, N (% col) | Low | 3,482 (65.2%) | 1,789 (60.4%) | 1,693 (71.1%) |
|  | Medium | 968 (18.1%) | 612 (20.7%) | 356 (14.9%) |
|  | High | 891 (16.7%) | 559 (18.9%) | 333 (14.0%) |
| Exposure to smoke at work, N (% col) | Yes | 1,081 (20.2%) | 574 (19.4%) | 508 (21.3%) |

N- number. % Col- column percentages. SD- standard deviation. Adjusted for sample weight, stratification and clustering.

### Model Performance Comparison

The performance metrics of the various machine learning models employed are summarized in Table 2. The Gradient Boosting Classifier demonstrated the highest accuracy (0.7478), AUC (0.7809), and F1 score (0.5798) among all the models, indicating its superior ability to predict multimorbidity in this population. Linear Discriminant Analysis and AdaBoost Classifier also exhibited strong performance, with AUC scores of 0.7777 and 0.7775, respectively.

**Table 2:** Model predictions comparison.

| Model | Accuracy | AUC | Recall | Prec. | F1 | Kappa | MCC |
| --- | --- | --- | --- | --- | --- | --- | --- |
| Gradient Boosting Classifier | 0.7478 | 0.7809 | 0.5132 | 0.6697 | 0.5798 | 0.4042 | 0.4124 |
| Linear Discriminant Analysis | 0.7430 | 0.7777 | 0.4935 | 0.6641 | 0.5654 | 0.3886 | 0.3978 |
| Ada Boost Classifier | 0.7418 | 0.7775 | 0.4956 | 0.6609 | 0.5653 | 0.3871 | 0.396 |
| Logistic Regression | 0.7407 | 0.7770 | 0.4766 | 0.6656 | 0.5546 | 0.3788 | 0.3898 |
| Ridge Classifier | 0.7400 | 0.7778 | 0.4759 | 0.6644 | 0.5536 | 0.3773 | 0.3883 |
| CatBoost Classifier | 0.7375 | 0.7742 | 0.5084 | 0.6463 | 0.5677 | 0.3832 | 0.3898 |
| Random Forest Classifier | 0.7290 | 0.76020 | 0.4671 | 0.6379 | 0.5383 | 0.3532 | 0.3623 |
| Light Gradient Boosting Machine | 0.7260 | 0.7578 | 0.4936 | 0.6229 | 0.55 | 0.3568 | 0.3623 |
| Extra Trees Classifier | 0.7200 | 0.7399 | 0.4759 | 0.6141 | 0.5357 | 0.3398 | 0.3459 |
| Naive Bayes | 0.7115 | 0.7465 | 0.5979 | 0.5716 | 0.5841 | 0.3635 | 0.364 |
| Quadratic Discriminant Analysis | 0.7085 | 0.7023 | 0.5124 | 0.5753 | 0.5313 | 0.3259 | 0.3309 |
| Extreme Gradient Boosting | 0.7085 | 0.7377 | 0.4874 | 0.5843 | 0.5308 | 0.3221 | 0.3252 |
| K Neighbors Classifier | 0.6818 | 0.6913 | 0.4258 | 0.5396 | 0.4752 | 0.2518 | 0.2558 |
| Dummy Classifier | 0.6609 | 0.5000 | 0 | 0 | 0 | 0 | 0 |
| Decision Tree Classifier | 0.6513 | 0.6132 | 0.4948 | 0.4862 | 0.49 | 0.2253 | 0.2255 |
| SVM - Linear Kernel | 0.6315 | 0.7051 | 0.4736 | 0.534 | 0.38 | 0.1789 | 0.2233 |

The Dummy Classifier, which served as a baseline model, achieved an accuracy of 0.6609 and an AUC of 0.5, confirming that all the employed models performed substantially better than random guessing. This finding underscores the value of using machine learning techniques to identify individuals at risk of multimorbidity based on their sociodemographic and health-related characteristics.

The confusion matrix (Figure 1) provides a more detailed breakdown of the Gradient Boosting Classifier’s performance. The model correctly identified 55.3% of the true positives (individuals with multimorbidity) and 89.2% of the true negatives (individuals without multimorbidity), demonstrating its ability to accurately classify a significant proportion of the study participants. However, the model misclassified 10.8% of the individuals with multimorbidity as not having the condition (false negatives) and 44.7% of the individuals without multimorbidity as having the condition (false positives), highlighting the need for further refinement of the model to improve its predictive accuracy.

**Figure 1:**
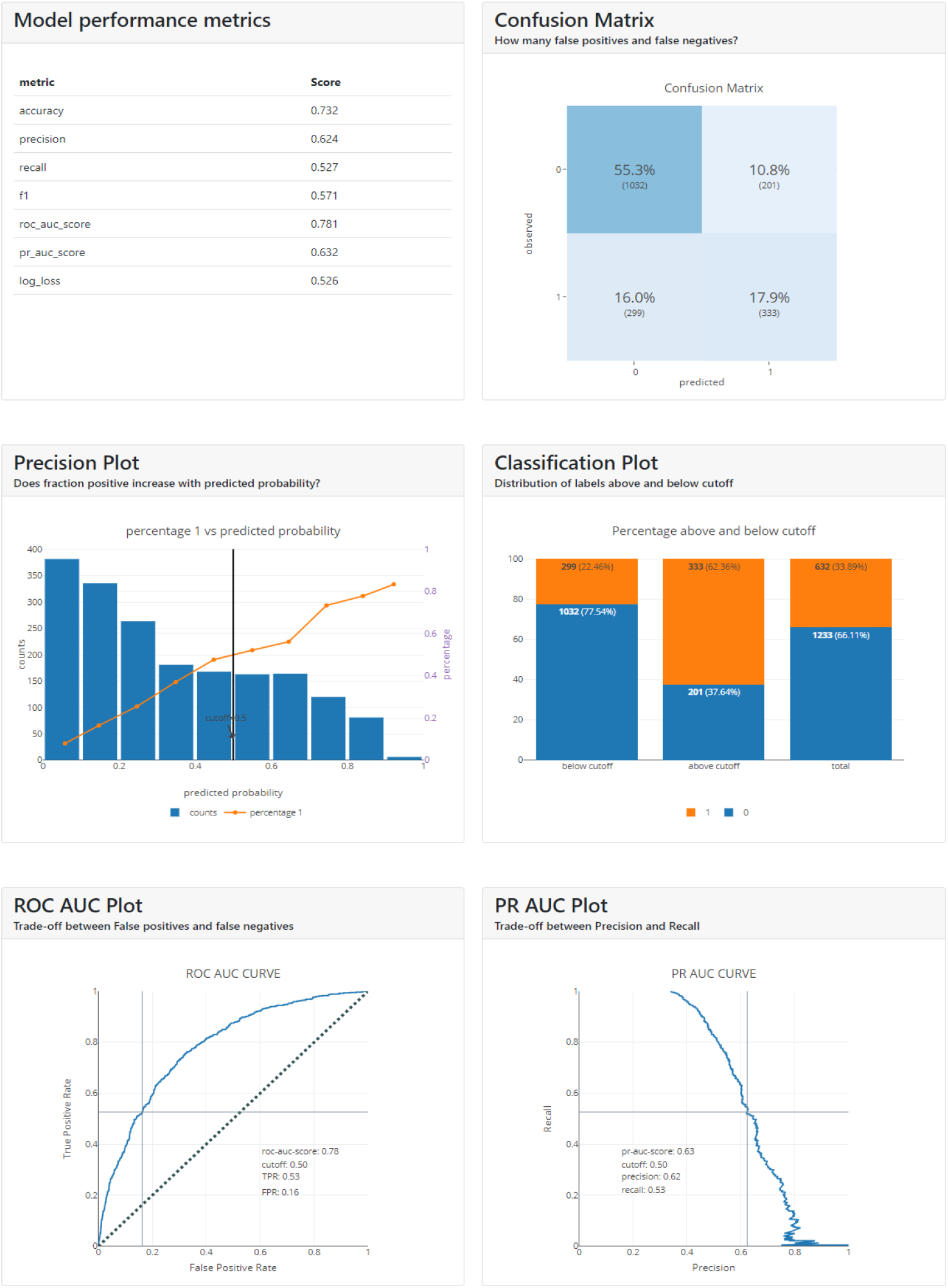
Summary metrics for best performing model.

### Feature Importance

The SHAP (Shapley Additive Explanations) values, depicted in Figure 2, provide insights into the average impact of the predictor variables on the model output. Age emerged as the most influential factor, followed by no medication use, female gender, poor health perception, and community illiteracy rate. These findings suggest that individual-level factors, such as age, gender, and health status, exert a more substantial influence on the likelihood of multimorbidity compared to community-level factors, such as illiteracy and poverty rates.

**Figure 2:**
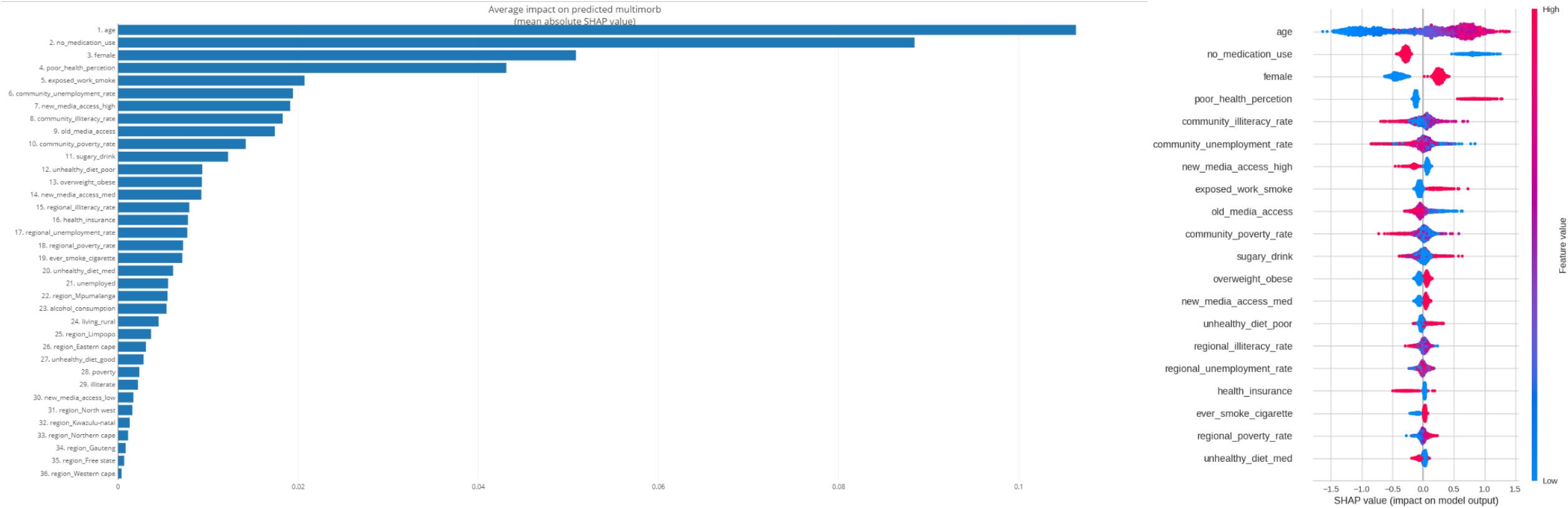
Variable importance.

### Individual-Level Feature Impact

A more granular analysis of the impact of individual-level features on the model output is provided in Figure 3. Higher values of age, female gender, poor health perception, no medication use, and overweight/obesity status were found to be associated with an increased likelihood of multimorbidity. These findings are consistent with previous research highlighting the role of aging, gender disparities, and modifiable risk factors, such as obesity and medication non-adherence, in the development of multiple chronic conditions. Conversely, younger age, being a male, no medication, good health perception were associated with a decreased likelihood of multimorbidity.

**Figure 3:**
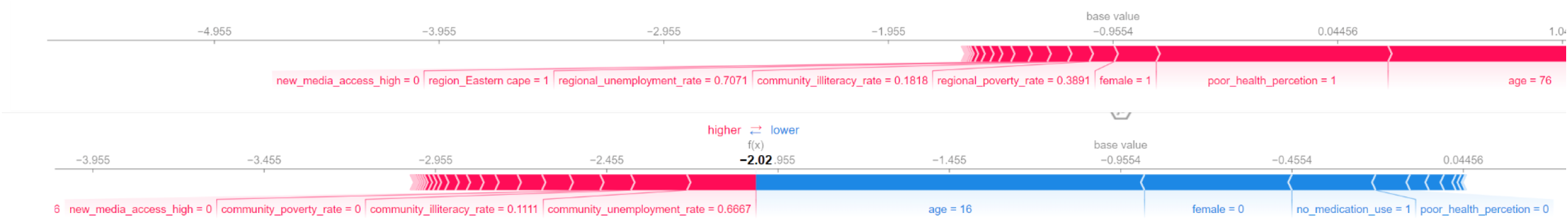
Shapley Addictive explanation (SHAP) force plot for two selected patients.

### Fairness and Demographic Bias Assessment

As shown in Figure 4A, the assessment of the model’s performance across different regions (Eastern Cape, Free State, Gauteng, KwaZulu-Natal, Limpopo, Mpumalanga, North West, Northern Cape, and Western Cape) revealed consistent results for all metrics, suggesting that the model is fair and unbiased with respect to the region feature. Similarly, the model maintained high levels of fairness across different wealth index categories (poorest to richest) for most metrics, with only slight variations in the Kappa and MCC metrics, which were marginally higher for the richest category.

**Figure 4:**
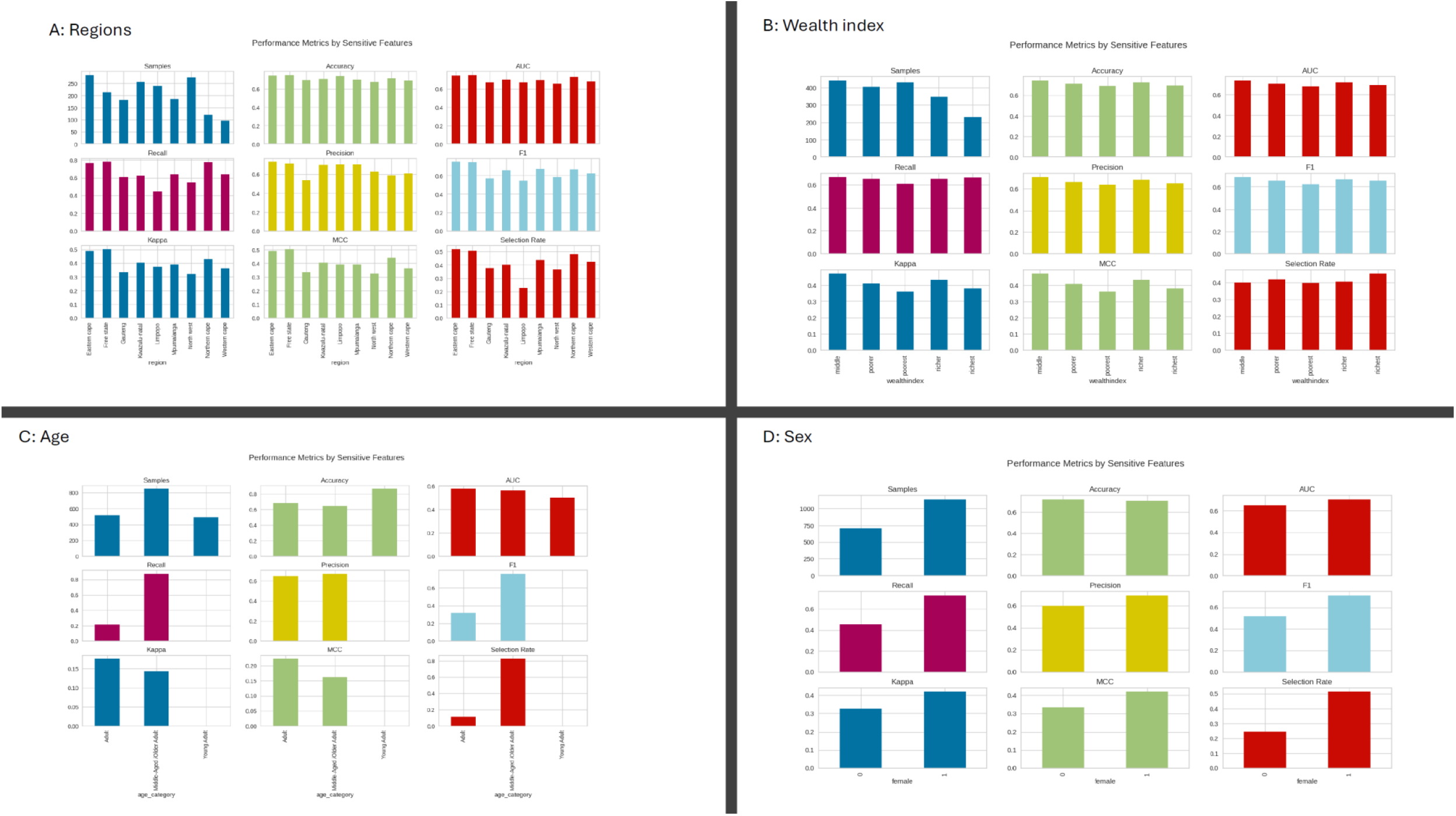
Fairness and Demographic Bias Assessment results.

Figure 4B compares the model’s performance across different wealth index categories, ranging from poorest to richest. The samples, precision, accuracy, recall, F1, and Selection Rate metrics are nearly identical across all wealth index categories, indicating a high level of fairness. However, there are slight variations in the Kappa metric, with the richest category having a marginally higher value compared to the other categories. This suggests that the model has a slightly better class agreement for the richest wealth index category. The MCC metric is also marginally higher for the richest category, indicating a slightly better overall performance. While these differences are small, they should be taken into account when interpreting the model’s results for each wealth index category. Further analysis could help identify the underlying reasons for these variations and ensure complete fairness across different socioeconomic groups.

Figure 4C analyses the model’s performance across four age categories: Adult, Middle-Aged, Older, and Young Adult. The model maintains high levels of fairness across all age groups for the samples, precision, accuracy, F1, and Selection Rate metrics. However, there are some variations in the Kappa, recall, and MCC metrics. The Young Adult and Adult categories have slightly higher Kappa and recall values compared to the Middle-Aged and Older categories. This indicates that the model might be slightly better at correctly identifying positive cases for younger age groups. The differences in MCC suggest that the model’s overall performance is somewhat better for younger age categories. While these variations are not drastic, they should be considered when applying the model to different age groups.

Figure 4D compares the model’s performance between the female and male genders. The samples, precision, accuracy, recall, F1, and Selection Rate metrics are nearly identical for both genders, indicating a high level of fairness. However, the Kappa metric is slightly higher for females, suggesting that the model has a better class agreement for the female gender. The MCC metric is also marginally higher for females, indicating a slightly better overall performance. While these differences are small, they should be taken into account when interpreting the model’s results for each gender. Further analysis could help identify the underlying reasons for these variations and ensure complete gender fairness.

## Discussion

### Main Findings

This study aimed to investigate the predictive performance of various machine learning models in identifying individuals at risk of multimorbidity in South Africa. The results revealed that the Gradient Boosting Classifier achieved the highest predictive performance, with an AUC of 0.7809, accuracy of 0.7478, and F1 score of 0.5798. The SHAP analysis identified age, no medication use, female gender, poor health perception, and community illiteracy rate as the most influential predictors of multimorbidity. Individual-level factors were found to have a more substantial impact on the likelihood of multimorbidity compared to community-level factors. However, the study also highlighted the importance of considering contextual factors, as higher community illiteracy rates and regional unemployment rates were associated with an increased likelihood of multimorbidity.

The fairness and demographic bias assessment revealed that the Gradient Boosting Classifier maintained a high level of fairness across different regions, wealth index categories, age groups, and genders. However, slight variations were observed in certain metrics for specific demographic categories. The model performed consistently across all regions, while the Kappa and MCC metrics were marginally higher for the richest wealth index category. The Young Adult and Adult age categories had slightly higher Kappa and recall values compared to the Middle-Aged and Older categories, and the Kappa and MCC metrics were slightly higher for females.

These findings suggest that an accurate classification algorithm, such as the Gradient Boosting Classifier, could serve as a clinically useful tool to advance the prevention and control of multimorbidity by empowering healthcare providers to prescribe preventative measures to at-risk patients and take earlier action to order diagnostic tests (Alonso-Morán et al., 2015). By identifying individuals at high risk of developing multiple chronic conditions, healthcare systems can allocate resources more effectively and implement targeted interventions to improve health outcomes and reduce the burden of multimorbidity (Arokiasamy et al., 2015). The study findings underscore the importance of considering both individual-level factors, such as age, gender, and health status, and community-level factors, such as illiteracy and unemployment rates, in understanding the determinants of multimorbidity in the South African context.

Higher values of community illiteracy rate and regional unemployment rate were found to be associated with an increased likelihood of multimorbidity, underscoring the potential influence of socioeconomic factors on health outcomes at the community level. These findings highlight the need for interventions that address the social determinants of health, such as education and employment, to reduce the burden of multimorbidity in disadvantaged communities.

Interestingly, higher values of age category and female gender were associated with a decreased likelihood of multimorbidity at the community level, in contrast to the individual-level findings. This observation may reflect the complex interplay between individual and contextual factors in shaping the risk of multiple chronic conditions and warrants further investigation to unravel the underlying mechanisms and potential effect modifiers.

These insights can inform the development of targeted interventions and policies aimed at preventing and managing multimorbidity, with a focus on addressing modifiable risk factors at the individual level and social determinants of health at the community level. Further research is needed to validate these findings in other populations and settings, and to explore the potential of integrating machine learning techniques into clinical decision support systems to improve the early detection and management of multimorbidity.

### Comparison with Previous Studies

The findings of this study are consistent with previous research highlighting the role of individual-level factors, such as age, gender, and health status, in the development of multimorbidity (Barnett et al., 2012; Marengoni et al., 2011). The higher prevalence of multimorbidity among females observed in this study aligns with the findings of a systematic review by Violan et al. (2014), which reported a consistent association between female gender and multimorbidity across various settings. The identification of poor health perception as a significant predictor of multimorbidity is supported by a study by Mavaddat et al. (2014), which found that self-rated health was strongly associated with the presence of multiple chronic conditions.

The importance of contextual factors, such as community illiteracy rates and regional unemployment rates, in predicting multimorbidity is consistent with the growing body of evidence on the social determinants of health (Marmot et al., 2008). A study by Nielsen et al. (2019) found that living in socioeconomically deprived areas was associated with an increased risk of multimorbidity, independent of individual-level factors. The findings of this study extend this line of research by demonstrating the potential of machine learning techniques to capture the complex interplay between individual and contextual factors in shaping the risk of multimorbidity.

### Implications for Practice and Future Research

The results of this study have important implications for clinical practice and public health policy. The accurate identification of individuals at high risk of multimorbidity using machine learning algorithms can facilitate the development of targeted interventions and care coordination strategies to prevent the onset and progression of multiple chronic conditions (Nicholson et al., 2019). Healthcare providers can use these tools to prioritize preventive care, such as lifestyle modifications and screening programs, for at-risk patients, potentially reducing the burden of multimorbidity and improving health outcomes (Salisbury et al., 2018).

Furthermore, the study highlights the need for a comprehensive approach to addressing the social determinants of health to reduce the burden of multimorbidity in disadvantaged communities. Public health policies and interventions should focus on improving access to education, employment opportunities, and healthcare services in these communities to mitigate the impact of socioeconomic disparities on health outcomes (Marmot et al., 2008). Future research should explore the effectiveness of community-level interventions in reducing the prevalence of multimorbidity and investigate the potential of integrating machine learning algorithms into clinical decision support systems to optimize the prevention and management of multiple chronic conditions (Nicholson et al., 2019).

### Study Strengths and Limitations

One of the main strengths of this study is the use of a large, nationally representative dataset, which enhances the generalizability of the findings to the South African population. The application of various machine learning algorithms and the rigorous evaluation of their predictive performance using multiple metrics provide a comprehensive assessment of the potential utility of these tools in identifying individuals at risk of multimorbidity. The use of SHAP values for model interpretation offers a transparent and interpretable approach to understanding the contribution of individual predictors to the model output, facilitating the translation of these findings into clinical practice (Lundberg & Lee, 2017).

However, the study also has several limitations that should be considered when interpreting the results. First, the cross-sectional design of the study precludes the establishment of causal relationships between the predictor variables and multimorbidity. Future research should employ longitudinal designs to investigate the temporal dynamics of these associations and to validate the predictive performance of the machine learning models over time. Second, the study relied on self-reported data for some of the predictor variables, such as health perception and medication use, which may be subject to recall bias and social desirability bias. Future studies should incorporate objective measures of health status and medication adherence to improve the accuracy of the predictive models.

## Conclusion

In conclusion, this study demonstrates the potential of machine learning algorithms, particularly the Gradient Boosting Classifier, in predicting multimorbidity in the South African context. The findings highlight the importance of considering both individual-level factors and contextual factors in understanding the determinants of multimorbidity. The accurate identification of individuals at high risk of developing multiple chronic conditions using these tools can inform the development of targeted interventions and care coordination strategies to prevent the onset and progression of multimorbidity, ultimately improving health outcomes and reducing the burden on healthcare systems. Future research should focus on validating these findings in other populations and settings, ideally in longitudinal studies, exploring the effectiveness of community-level interventions in reducing the prevalence of multimorbidity, and investigating the potential of integrating machine learning algorithms into clinical decision support systems to optimize the prevention and management of multiple chronic conditions, in randomised controlled trials.

## Data Availability

All data produced in the present work are contained in the manuscript

## Notes

### Competing Interest Statement

The authors have declared no competing interest.

### Funding Statement

This study did not receive any funding

